# CSF TDP-43: A Novel Biomarker for Limbic-Predominant Age-Related TDP-43 Encephalopathy

**DOI:** 10.64898/2026.05.07.26352671

**Authors:** Anna-Lisa Fischer, Franziska Flosbach, Elisabeth Root, Maren Breitbarth, Bettina Goericke, Matthias Schmitz, Peter Hermann, Inga Zerr

## Abstract

Mislocalization and aggregation of transactive response DNA-binding protein 43 kDa (TDP-43) represent a neuropathological hallmark of amyotrophic lateral sclerosis (ALS) and frontotemporal lobar degeneration (FTLD) and are increasingly recognized in Alzheimer’s disease (AD) and limbic-predominant age-related TDP-43 encephalopathy (LATE). However, the in vivo value of CSF TDP-43 as a biomarker and its relation to established markers remains unclear. We quantified CSF concentrations of TDP-43 using ELISA in 25 controls, 32 ALS, 9 probable LATE, and 24 AD patients. CSF TDP-43 levels differed significantly between groups, with the highest concentrations in LATE, exceeding both ALS and AD. ALS and AD showed intermediate, comparable increases versus controls. In parallel, conventional AD biomarkers (t-tau, p-tau, and amyloid-b) showed the expected AD-typical profile but remained largely unaltered in probable LATE, indicating a dissociation between TDP-43 an AD-type pathology. These findings identify CSF TDP-43 as a promising candidate biomarker for LATE, characterized by disproportionate elevation in the absence of AD-type biomarker changes, and neurodegeneration in aging populations.

## Introduction

Contemporary global demographics reveal an unprecedented expansion of population aged 80 years and beyond, presenting challenges of age-related neurodegeneration.^1^ Dementia prevalence demonstrates a steep age-dependent gradient, forecasting a substantial increase in overall dementia burden. Emerging evidence has identified previously unrecognized neuropathological entities, such as limbic-predominant age-related TDP-43 encephalopathy neuropathologic change (LATE-NC), which accounts for a substantial proportion of patients which present with amnestic dementia.^1-4^ The disease is defined by neuropathological abnormalities and clinical criteria have been suggested. However, no disease-specific biomarker has been identified so far to support the clinical diagnosis.

Transactive response DNA-binding protein 43 kDa (TDP-43) is a nuclear RNA and DNA-binding protein with essential roles in RNA metabolism, whose pathological mislocalization and aggregation constitute a molecular hallmark of amyotrophic lateral sclerosis (ALS) and frontotemporal lobar degeneration (FTLD).^5^ Increasing evidence indicates that TDP-43 pathology extends beyond these classical TDP-43 proteinopathies, being detected in up to 50% of Alzheimer’s disease (AD) cases and defining the recently characterized LATE.^3,6^ In AD, TDP-43 pathology correlates with faster cognitive decline, whereas in LATE it represents the primary neuropathological substrate, often mimicking the clinical presentation of AD but with distinct neuroanatomical predilections.^7^

Despite extensive post-mortem evidence, the in vivo quantification of TDP-43 remains elusive. Reported cerebrospinal fluid (CSF) concentrations vary across studies, reflecting methodological heterogeneity, preanalytical variability, and uncertainty about whether soluble TDP-43 species reliably track neuropathological burden. Few studies have directly compared CSF TDP-43 across multiple neurodegenerative diseases within the same analytical framework, and almost none have systematically integrated these measurements with established CSF biomarkers – total tau (t-tau), phosphorylated tau (p-tau), and amyloid-β (Aβ), to evaluate their relative or complementary diagnostic value. Importantly, no data are available for patients with LATE.

LATE poses a particular diagnostic challenge and remains under-recognized in clinical practice. Here, we quantified CSF TDP-43 by enzyme-linked immunosorbent assay (ELISA) in well-characterized cohorts of controls, ALS, AD, and LATE patients. We hypothesized that TDP-43 concentrations would differ substantially between these disorders, and that LATE would display a distinct biomarker signature compared with both ALS and AD. We further examined correlations with t-tau, p-tau, and Aβ to explore the potential of TDP-43 as a part of a multimarker approach to differential diagnosis and defined the CSF protein profile in LATE.

## Materials and methods

### Patients

The study recruited 94 patients. All participants were enrolled at the Clinical Dementia Center at the National Reference Center for Transmissible Spongiform Encephalopathies, located within the Department of Neurology at the University Medical Center Goettingen, Germany.

### Diagnostic criteria

#### Controls and ALS

Control (CTRL) participants were recruited from patients presenting with non-neurodegenerative conditions. The primary diagnoses in this group included psychiatric disorders, headache syndromes, peripheral neuropathies, syncope, vertigo (attributed to medication side effects), and benign intracranial hypertension. None of the control individuals showed clinical or diagnostic evidence of a neurodegenerative disease. ALS patients were diagnosed according to revise El Escorial clinical criteria for ALS.

#### LATE

The diagnosis of LATE was established using a multimodal clinical classification framework integrating CSF biomarkers, neuropsychological assessment and structural neuroimaging, in accordance with current clinicopathological concepts of LATE.^8-9^ Standardized neuropsychological assessment focused on episodic memory performance, as LATE is characteristically associated with a predominantly amnestic cognitive syndrome. ^9^ Then, CSF biomarkers were used to stratify patients with amnestic clinical syndrome by AD pathology. Patients with negative amyloid and tau biomarkers were classified as probable LATE (A-/T-/N+), indicating a CSF profile inconsistent with AD.^10^ Patients with marked episodic memory deficits, with or without additional impairments in semantic fluency or working memory, were considered compatible with a LATE phenotype.^11-14^ Structural MRI was evaluated and pronounced medial temporal lobe atrophy, particularly involving the hippocampus, was considered supportive of LATE, consistent with the known of TDP-43 pathology.^11^ Patients with at least mild cognitive impairment or dementia were classified as LATE-suspect cases when they fulfilled biomarker constellations compatible with probable LATE or possible LATE and showed predominantly amnestic clinical phenotype.

#### AD

All patients in the AD group fulfilled clinical criteria for Alzheimer’s dementia and CSF analyses showed positivity of AD-related amyloid and tau pathology (A+/T+).^10^ Concomitant CNS pathologies such as significant vascular injury or other causes of dementia syndromes were clinically excluded as far as possible through standard diagnostic work-up including MRI, lumbar puncture, and clinical examination.

### CSF biomarkers collection and analysis

CSF was collected for diagnostic purposes. After the routine analyses, remained CSF was stored at – 80°C. TDP-43 concentrations were measured using the human TDP-43 ELISA kit (Proteintech, Chicago, USA) following the manufactures instructions. CSF samples were diluted 1:2 prior to analysis. The limit of quantification was 6.25 ng/ml, and the limit of detection was 0.26 ng/ml. CSF concentrations of t-tau, p-tau, Ab42, and Ab40 were quantified using Lumipulse kits (Fujirebio, Ghent, Belgium) by the neurochemistry laboratory at department of Neurology, University Medical Center Goettingen, using standard procedures. Pathological thresholds were determined based on established laboratory-specific cut-offs: t-tau >449 pg/mL, p-tau >60 pg/mL, Abeta 1-42 <450 pg/mL, and Abeta ratio <0.5.

### Statistical analysis

To compare TDP-43 levels between groups, data were first tested for normality. Group comparisons were performed using either one-way ANOVA or the non-parametric Kruskal-Wallis test, as appropriate. For the assessment of significant differences in CSF levels, including TDP43, t-tau, p-tau, Ab40 and Ab42, group comparisons were conducted using the Kruskal-Wallis test followed by uncorrected Dunn’s post hoc test. Differences between two independent groups were analyzed using Mann-Whitney U test. Correlations between variables were assed using Spearman’s rank correlation coefficient. All statistical analyses were performed using GraphPad Prism version 10.1.2.

## Results

### Clinical data

Table 1 presents demographic characteristics and CSF biomarker profiles across four clinical groups: CTRL, ALS, probable LATE, and AD patients. The cohort comprised 94 patients, with CTRL (*n*=29) being younger (mean age 49.5±17.3) and showing a balanced sex distribution (13 females/ 16 males). The ALS group (*n*=32) showed intermediate age (64.2±13.3) and similar sex distribution (14/18), while both AD (*n*=24) and LATE (*n*=9) were elderly groups with comparable mean ages (76.8±8.1 and 78.5±5.3 years, respectively). The key finding in Table 1 is markedly elevated CSF TDP-43 concentration in LATE group (24.41±5.36 pg/ml), 2.3-fold higher than ALS (10.8±7.6 pg/ml), and 1.9-fold higher than AD (13.2±4.1 pg/ml). Conventional AD biomarkers showed expected patterns: t-tau and p-tau were most elevated in AD patients (738.4±283.9 pg/ml and 113.8±30.6 pg/ml, respectively), while Ab42/Ab40 ratio was characteristically reduced in AD (0.4±0.1) compared to controls (1.3±0.3). The LATE group exhibited t-tau and p-tau levels within the range of CTRL and ALS and largely preserved Ab42/Ab40 ratio (1.15±0.23), suggesting limited contribution of AD-type biomarker alterations despite the advanced age of this cohort. Routine CSF parameters were mainly unremarkable across groups. Qalb values and total CSF protein levels were within age-appropriate reference limits, and CSF lactate remained in the normal range. These findings indicate that group differences in CSF TDP-43 are unlikely to be driven by overt blood-CSF barrier dysfunction or major metabolic disturbance.

**Table 1:** Demographic data and biomarker concentrations.

|  | CTRL | ALS | probable LATE | AD |
| --- | --- | --- | --- | --- |
| **Age (mean ± SD)** | 49.5 ± 17.3 | 64.2 ± 13.3 | 78.5 ± 5.3 | 76.8 ± 8.1 |
| **Sex (f/m)** | 13/16 | 14/18 | 4/5 | 13/11 |
| **TDP-43 (pg/ml)** | 9.1 ± 10.5 | 10.8 ± 7.6 | 24.41 ± 5.36 | 13.2 ± 4.1 |
| **t-tau (pg/ml)** | 248.1 ± 87.6 | 328.3 ± 151.5 | 260.4 ± 130.0 | 738.4 ± 283.9 |
| **p-tau (pg/ml)** | 44.5 ± 15.9 | 54.1 ± 22.3 | 50.9 ± 16.2 | 113.8 ± 30.6 |
| **Ab42 (pg/ml)** | 1232.6 ± 465.1 | 1302.5 ± 400.9 | 1541.7 ± 565.4 | 525.1 ± 180.3 |
| **Ab40 (pg/ml)** | 9803.4 ± 3604.8 | 9977.7 ± 2067.7 | 13002 ± 3099 | 13060.5 ± 3389.8 |
| **Ab-ratio** | 1.3 ± 0.3 | 1.4 ± 0.5 | 1.15 ± 0.23 | 0.4 ± 0.1 |

Data are presented as mean ± standard deviation, except for sex. Abbreviations: CTRL, control; ALS, amyotrophic lateral sclerosis; LATE, Limbic-predominant Age-related TDP-43 Encephalopathy; AD, Alzheimer's disease; TDP-43, Transactive response DNA-binding protein 43 kDa; t-tau, total tau; p-tau, phosphorylated tau; Ab42, amyloid beta 42; Ab40, amyloid beta 40.

### ELISA

CSF TDP-43 levels were measured by ELISA in CTRL (*n*= 29), ALS (*n*=32), probable LATE (*n*=9), and AD (*n*=24) (Figure 1). Established biomarkers (t-tau, p-tau, Ab40, Ab42) were analyzed in parallel. CSF biomarker profiles differed in a disease-specific manner. CSF TDP-43 levels varied significantly (Kruskal-Wallis test, *P*<0.0001). Compared with CTRL, CSF TDP-43 concentrations were modestly increased in ALS (*P*=0.0235) and AD (*P*=0.0028), whereas patients with probable LATE showed a pronounced elevation (*P*<0.0001). Direct comparisons further showed higher CSF TDP-43 levels in probable LATE than in ALS (*P*=0.0014) and AD (*P*=0.0239), while no difference was observed between ALS and AD (*P*>0.9999), indicating that elevated CSF TDP-43 is not a general feature of neurodegeneration but is particularly associated with probable LATE.

**Figure 1.**
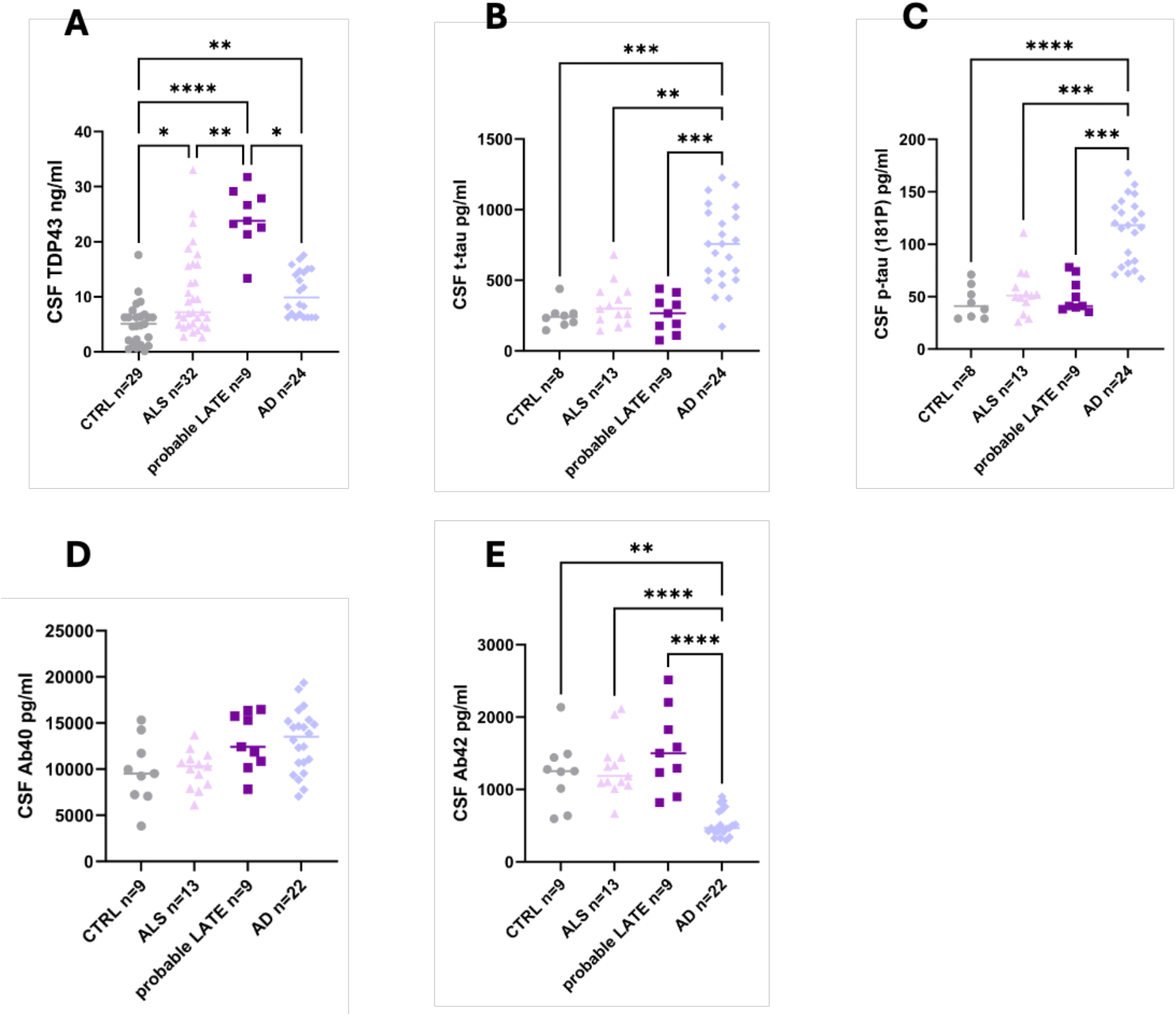
Distinct CSF biomarker profiles across neurodegenerative disease entities. Scatter plots illustrate CSF concentrations of TDP-43 (A), t-tau (B), p-tau (181) (C), Ab40 (D), and Ab42 (E) in CTRL, ALS, probable LATE, and AD patients as measured by ELISA or Lumipulse. Individual measurements are shown as dots, with horizontal lines indicating the mean for each group. CSF TDP-43 concentrations differed significantly between groups, with the highest levels observed in probable LATE, compared with CTRL, ALS and, AD. In contrast, AD patients exhibited the most pronounced elevations in CSF t-tau and p-tau, accompanied by a marked reduction of CSF Ab42, whereas these biomarkers remained largely preserved in probable LATE. No significant group differences were detected for CSF Ab40. Sample sizes vary across biomarkers, as analyses were restricted to subjects with available measurements for the respective analyte. Data are presented as mean SD. Statistical analysis was performed using a Kruskal-Wallis test followed by Dunn’s multiple comparisons test (^*\**^*P*<0.05, ^*\*\**^*P* < 0.01, ^*\*\*\**^*P* < 0.001, ^*\*\*\*\**^*P*< 0.0001).

In contrast, tau biomarkers clearly segregated AD from the other groups. CSF t-tau were increased in AD compared with CTRL (*P*=0.0004), ALS (*P*=0.0011), and probable LATE (*P*=0.0004), with no differences between CTRL, ALS, and probable LATE (all *P*>0.9999). A similar pattern was observed for p-tau, which was elevated in AD relative to CTRL (*P*<0.0001), whereas p-tau levels in probable LATE did not differ from CTRL or ALS (both *P*>0.9999). Amyloid biomarkers further supported a biochemical dissociation between AD and probable LATE. CSF Ab42 concentrations were reduced in AD compared with CTRL (*P* =0.0016), ALS (*P*<0.0001), and probable LATE (*P*<0.0001), no differences were observed between probable LATE and CTRL (*P*>0.9999) or ALS (*P*>0.9999). CSF Ab40 concentrations did not differ significantly between any of the groups. Taken together, these results demonstrate that LATE is characterized by disproportionately elevated CSF TDP-43 in the absence of tau- or amyloid-associated alterations, yielding a CSF biomarker profile that is distinct from both AD and ALS.

### Correlation analysis

Following the demonstration of distinct CSF TDP-43 profiles across diagnostic categories, we investigated the associations between CSF TDP-43, demographic variables, and established CSF biomarkers to delineate the biological correlates of elevated TDP-43 levels (Table 2). Within the probable LATE cohort, CSF TDP-43 showed no association with age (Spearman r=-0.15, *P*=0.70), indicating that elevated TDP-43 concentrations were not driven by age alone. CSF TDP-43 levels correlated moderately with t-tau (r=0.67, *P*=0.0499), whereas no significant associations were observed with p-tau (r=0.17, *P*=0.67) or the Ab40/42 ratio (r=0.47, *P*=0.20). In AD cohort, CSF TDP-43 did not correlate with t–tau (r=-0.13, *P*=0.63), p-tau (r=0.10, *P*=0.72), or the Ab42/40 ratio (r=0.29, *P*=0.27), and showed only a trend toward a negative association with age (r=-0.49, *P*=0.057). Similarly, in ALS, no associations were observed between CSF TDP-43 and age (r=-0.14, *P*=0.66), t-tau (r=-0.31, *P*=0.51), or p-tau (r=-0.56, *P*=0.059), although a significant positive correlation emerged between CSF TDP-43 and the Ab42/40 ratio (r=0.74, *P*=0.006). In controls, no significant correlations were detected between CSF TDP-43 and any of the investigated variables. Across the entire cohort, CSF TDP-43 did not show consistent correlations with amyloid biomarkers or p-tau, supporting the dissociation between TDP-43 and AD-specific biomarker pathways. Together, these findings suggest that elevated CSF TDP-43 in LATE is not primarily associated with amyloid deposition or tau phosphorylation but may partially relate to markers of neuronal injury.

**Table 2:** Correlation analysis between CSF TDP-43 and demographic and biomarker variables in different diagnostic groups.

| **group** | **Age** | **t-tau** | **p-tau** | **Ab-ratio** |
| --- | --- | --- | --- | --- |
| **CTRL (n=8)** | -0.071 (0.867) | 0.690 (0.058) | 0.587 (0.126) | -0.214 (0.610) |
| **ALS (n=12)** | -0.140 (0.664) | -0.210 (0.512) | -0.559 (0.059) | 0.744 (0.006) |
| **probable LATE (n=9)** | -0.150 (0.700) | 0.667 (0.050) | 0.167 (0.668) | -0.471 (0.201) |
| **AD (n=16)** | -0.485 (0.057) | -0.129 (0.633) | 0.097 (0.721) | 0.292 (0.273) |

Values represent Spearman's rank correlation coefficients (r) with corresponding p-values in parentheses. Correlation was calculated to assess associations between CSF TDP-43 concentrations and age, as well as tau and amyloid biomarkers, across CTRL, ALS, probable LATE, and AD cohorts. Analyses were restricted to subjects with complete paired measurements. Given the limited sample sizes within each group, findings should be regarded as exploratory.

## Discussion

In this study, we provide the first systematic comparison of CSF TDP-43 concentrations across well-characterized cohorts of CTRL, ALS, LATE, and AD patients using standardized ELISA methodology. The principal finding is a pronounced elevation of CSF TDP-43 in probable LATE, with concentrations exceeding those observed in ALS and AD by approximately twofold. Importantly, this increase occurred in the absence of concomitant tau or amyloid abnormalities, establishing a CSF biomarker profile that is distinct from AD and not explained by generalized neurodegeneration alone.

### CSF TDP-43 as a candidate biomarker for LATE

LATE is defined by neuropathological criteria, and its clinical diagnosis remains challenging because of substantial phenotypic overlap with AD. Despite compelling post-mortem evidence identifying TDP-43 pathology as the defining molecular substrate of LATE, no validated in vivo biomarker is available to support clinical recognition. Our findings address this gap by demonstrating markedly elevated CSF TDP-43 concentrations in probable LATE compared with both ALS and AD, suggesting that CSF TDP-43 may serve as a disease-relevant biochemical marker rather than a nonspecific indicator of neurodegeneration. The magnitude of CSF TDP-43 elevation in LATE is notable. While modest increases were detected in ALS and AD relative to control, only LATE exhibited a pronounced and consistent elevation. This pattern argues against the notion that increased CSF TDP-43 simply reflects neuronal loss or disease severity and instead supports a closer relationship with the underlying TDP-43 proteinopathy that defines LATE.

### Dissociation from AD biomarker pathways

A key observation of this study is the clear dissociation between CSF TDP-43 and established AD biomarkers. In contrast to AD, where elevated t-tau and p-tau and reduced Ab42 were consistently observed, probable LATE patients exhibited tau and amyloid biomarker levels largely within the range of controls and ALS. This finding is particularly relevant given the advanced age of the LATE cohort and frequent co-occurrence of AD-type pathology in elderly individuals. The absence of tau and amyloid abnormalities in probable LATE underscores the biological distinctiveness of this entity and supports the concept that LATE represents a primary TDP-43-driven neurodegenerative process rather than a variant of AD. From clinical perspective, this dissociation highlights the potential utility of CSF TDP-43 as part of a biomarker panel to improve differential diagnosis in older patients presenting with amnestic dementia.

### Interpretation of correlation analyses

Correlation analyses inform the biological interpretation of elevated CSF TDP-43. Within the probable LATE group, CSF TDP-43 levels was not associated with age, indicating that increased levels are not merely an epiphenomenon of aging. The lack of correlation with p-tau and Ab42/40 ratio reinforces the independence of CSF TDP-43 from AD-specific pathological pathways. CSF TDP-43 showed a moderate correlation with t-tau within the LATE cohort. Total tau is widely regarded as a marker of neuronal injury rather than a disease-specific process, suggesting that CSF TDP-43 may partially reflect ongoing neurodegeneration in the context of TDP-43 pathology. However, the absence of a corresponding association with p-tau argues against a direct link to tau phosphorylation or tangle pathology. Together, these findings support a model in which CSF TDP-43 reflects a disease-relevant process related to TDP-43-mediated neurodegeneration rather than secondary AD-type pathology.

### Context within the literature

To date, only a limited number of studies have investigated TDP-43 in CSF, and none have focused on LATE. As summarized in Table 3, previous investigations have primarily addressed ALS and FTLD, often reporting modest CSF TDP-43 elevations with substantial interindividual variability and limited diagnostic accuracy. Methodological heterogeneity, including differences in assay platforms, antibody specificity, and preanalytical handling, has complicated cross-study comparisons. More recent studies employing ultrasensitive or in-house assays have suggested associations between CSF (phosphorylated) TDP-43 and neuroimaging or vascular markers, particularly in AD and mixed dementia cohorts. However, these studies have not established CSF TDP-43 as a disease-specific biomarker, nor have they systematically compared TDP-43 across multiple neurodegenerative conditions within a single analytical framework. Recent advances beyond CSF biomarkers underscore the potential of TDP-43-related measures in vivo. The study by Wang and colleagues^15^ demonstrated that plasma total TDP-43 was significantly elevated in individuals with advanced LATE-NC and that both total and phosphorylated plasma TDP-43 correlated with post-mortem brain TDP-43 burden, particularly in the context of comorbid AD pathology. Although the focus was on plasma rather than CSF, these findings converge with our CSF data by supporting the notion that TDP-43 can be detected in biofluids and may reflect underlying neuropathological processes. Our study extends this literature by providing a direct, standardized comparison across ALS, AD, and LATE, thereby revealing a disease-specific CSF TDP-43 signal that has not been previously recognized.

**Table 3:**
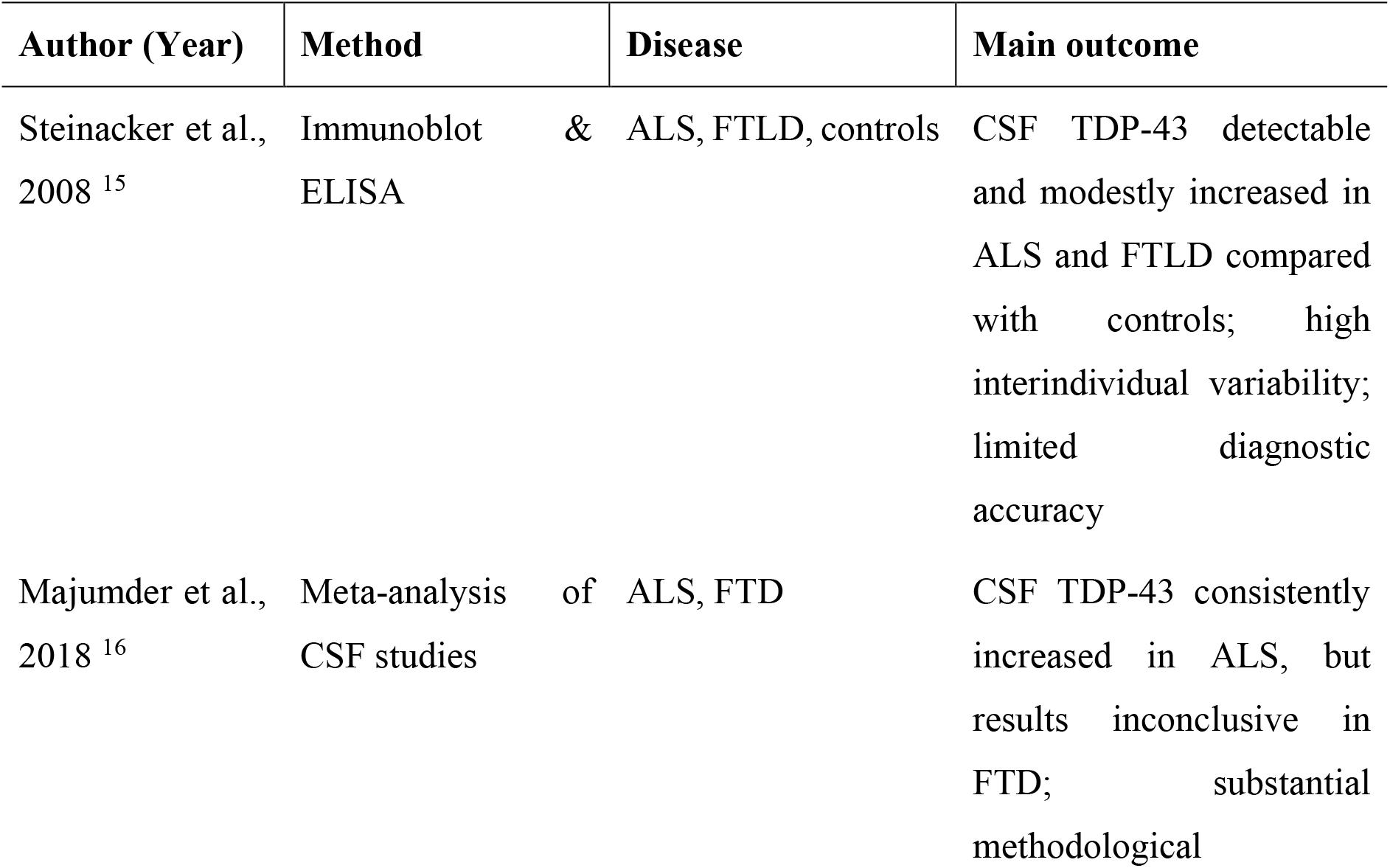
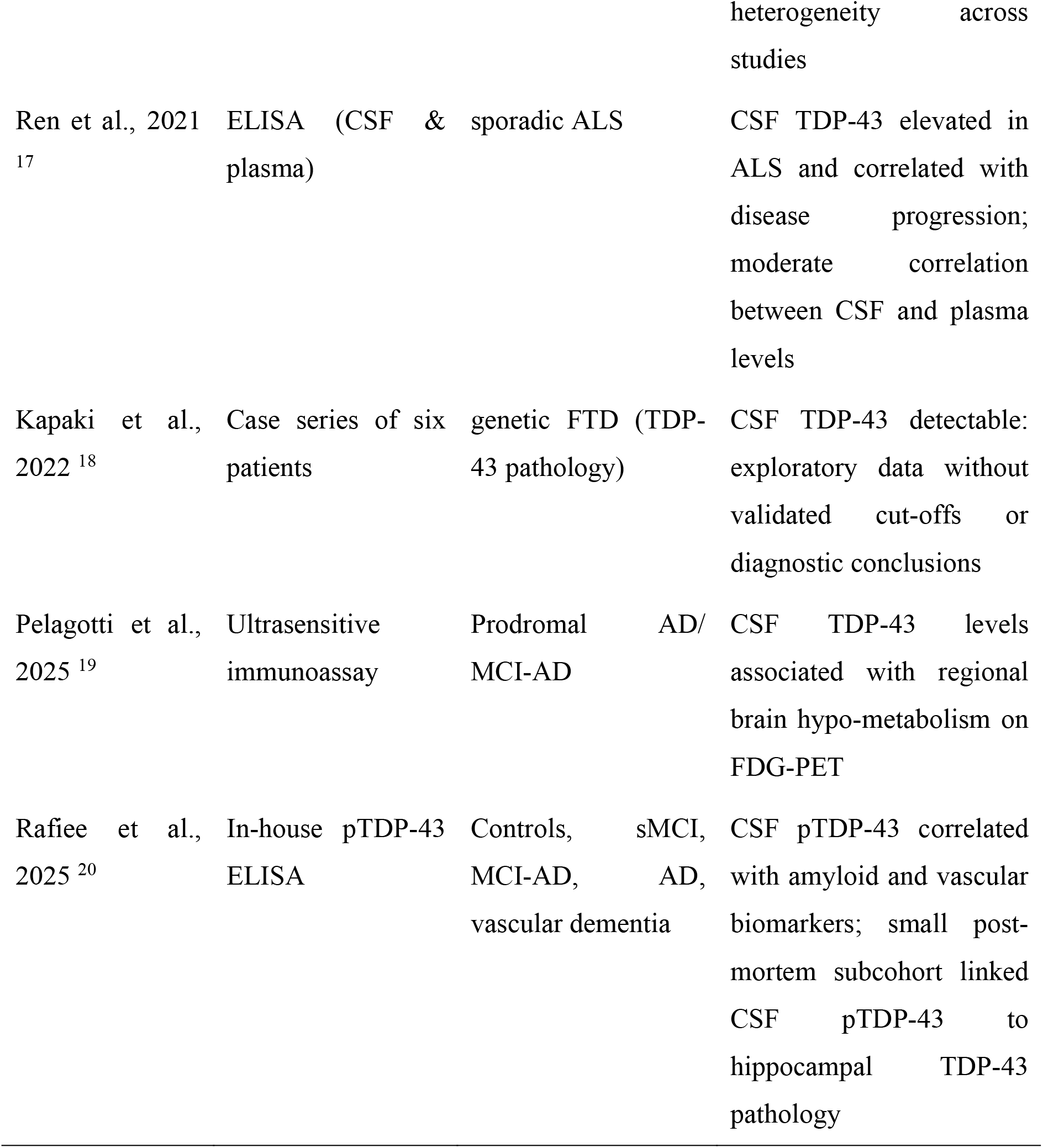
Overview of published studies assessing CSF TDP-43 in neurodegenerative diseases.

### Limitations and future directions

Several limitations warrant consideration. The probable LATE cohort was small, reflecting ongoing challenges in clinical identification in the absence of validated biomarkers. Accordingly, correlation analyses should be interpreted as exploratory. Neuropathological confirmation and longitudinal data were lacking. Future studies incorporating post-mortem validation, longitudinal sampling, and standardized assay platforms are required to define the temporal dynamics and diagnostic utility of CSF TDP-43.

In summary, CSF TDP-43 represents a promising biomarker for LATE, characterized by marked elevation in the absence of AD-type changes. The findings support its further evaluation for the in vivo characterization of TDP-43-associated neurodegeneration in aging populations.

## Data Availability

All data produced in the present study are available upon reasonable request to the authors

